# Synuclein and dopamine transporter biomarkers among phenoconverters to parkinsonian disorders

**DOI:** 10.64898/2026.04.15.26350768

**Authors:** Cristina Simonet, Jie Yin, Lana M. Chahine, Daniel Weintraub, Krittika Chatterjee, Chelsea Caspell-Garcia, David-Erick Lafontant, Alastair Noyce, Andrew Siderowf, Carlie Tanner, Ethan Brown, Thomas F. Tropea, Brit Mollenhauer, Roy N. Alcalay, Kathleen Poston, Kenneth Marek, Tanya Simuni, the Parkinson’s Progression Markers Initiative investigators

**Author notes:** See Supplementary Table_PPMI Contributors List Affiliations (the Parkinson’s Progression Markers Initiative investigators). Corresponding author: Tanya Simuni, MD Department of Neurology, Northwestern University Feinberg School of Medicine 710 North Lake Shore Drive, 1126, Chicago, IL 60611.

## Abstract

**Background:** Phenoconversion to Parkinson’s disease (PD) or dementia with Lewy bodies (DLB) currently relies on established clinical diagnostic criteria. Availability of *in vivo* biomarkers—CSF α-synuclein seed amplification assay (CSFaSynSAA) and dopamine transporter (DAT) imaging—offer the opportunity to investigate congruency between clinical phenoconversion and biologically defined disease.

**Methods:** We analyzed Parkinsońs Progression Markers Initiative participants who phenoconverted to PD, DLB, multiple system atrophy (MSA), Alzheimer’s disease (AD) or other dementias from prodromal and non-manifesting genetic carrier (NMC) groups and controls. Site investigators determined phenoconversion based on established diagnostic criteria. All phenoconverters with ≥1 annual follow-up visit, with available biomarkers and persistent clinically defined diagnosis at last observation were included. Neuronal alpha-Synuclein Disease Integrated Staging System (NSD-ISS) staging was applied.

**Results:** Among 121 phenoconverters, 103 had evaluable CSFaSynSAA and DAT data and were included in analysis: 92 PD, 7 DLB, 2 MSA, 2 AD/other dementias. Phenoconversion annual rates varied widely across groups: iRBD 7.9%, hyposmia 4.2%, *GBA1* 0.3%, *LRRK2* 1.3%, *LRRK2*+*GBA1* 0.9%, and controls 0.5%. Median time from baseline to phenoconversion ranged from 13-14 months in iRBD and hyposmia to 36-85 months in NMCs. The expected biomarker profile (CSFaSynSAA+/DAT+) for clinically-diagnosed synucleinopathy occurred in 74 (71.8%) participants. Biological alignment (CSFaSynSAA+/DAT+) was present in 87% hyposmics and 72% iRBD phenoconverters. CSFaSynSAA negativity was high among *LRRK2* phenoconverters (67%), who also were more likely to have a preserved sense of smell (83%). Phenoconversion occurred later than onset of functional impairment: 15/47 (31.9%) iRBDs and 7/38 (18.4%) hyposmics were already NSD-ISS stage ≥4 at time of phenoconversion.

**Conclusions:** Clinical phenoconversion did not necessarily align with biological evidence of synucleinopathy or dopaminergic loss and can be delayed compared to onset of meaningful functional impairment. Longitudinal follow up on converters without biological evidence of PD is required to confirm conversion diagnosis and evaluate for a later occurrence of biomarker positivity.

## Introduction

Reliable assessment of the transition from a prodromal state to clinically defined Parkinson’s disease (PD), dementia with Lewy bodies (DLB), and other related neurodegenerative conditions (e.g., multiple system atrophy (MSA), and progressive supranuclear palsy (PSP)), is essential for early detection and disease-modifying trials. The concept of phenoconversion, broadly defined as the emergence of clinical signs sufficient for a syndromic diagnosis, is an essential milestone in the prodromal population.

Phenoconversion has typically been treated as a binary event marking the point at which motor or cognitive symptoms meet threshold for clinical diagnostic criteria. However, binary clinical phenoconversion may fail to capture the temporal onset or magnitude of underlying pathophysiology. Progression is on a continuum and operationalizing phenoconversion as an endpoint for evolving disease-modifying clinical trials is challenging. In addition, accuracy of the clinically-defined diagnosis against ultimate pathological confirmation is the lowest in early clinical stages of PD.^1^ The 2015 Movement Disorder Society research criteria for prodromal PD and the 2017 DLB Consortium criteria both acknowledge these challenges and propose the integration of indicative biomarkers to improve diagnostic certainty.^2–4^

Recent advances in molecular and imaging biomarkers, particularly CSF α-synuclein Seed Amplification Assays (CSFaSynSAA) for abnormal aggregation of α-synuclein and dopamine transporter SPECT scan (DAT) imaging for nigrostriatal integrity, allow to accurately define *in vivo* disease biology years before an individual reaches the stage of clinically-defined diagnosis.^5,6^ Two biological diagnostic frameworks (“Synucleinopathy-Neurodegeneration-Genetics” (SynNeurGe) and “Neuronal α-Synuclein Disease” (NSD)) have been proposed.^7,8^ These frameworks revisit biological and clinical progression of PD on a continuum as an alternative to the canonical phenoconversion by incorporating *in vivo* biological characterization of individuals.

In many cases, clinical diagnosis aligns with underlying synucleinopathy and dopaminergic neurodegeneration. We refer to this alignment as biological congruence, defined as the presence of CSFaSynSAA together with dopaminergic deficit on DAT imaging in individuals carrying a clinical PD diagnosis. In two large longitudinal cohorts, a high proportion of people with sporadic PD were CSFaSynSAA+: 87% in the Parkinson’s Progression Markers Initiative (PPMI) study and 96% in the UK parkinsonism cohort.^9,10^ However, congruence varies in genetic PD like *LRRK2* variant carriers, as current CSFaSynSAA assays may not necessarily detect all pathogenic α-synuclein conformations.^11^

The clinically-defined prodromal state represents a heterogeneous group of individuals such as those with isolated REM sleep behavior disorder (iRBD), hyposmia, and non-manifesting carriers (NMCs) of PD-associated genetic variants (*GBA1, LRRK2, SNCA*); substantial variability exists in penetrance, phenotype, and biomarker expression.

Therefore, a key unanswered question is whether phenoconversion, defined by investigator-assigned clinical diagnosis, accurately reflects the biological state of individuals as determined by CSFaSynSAA and DAT imaging. Equally important is whether the timing of phenoconversion corresponds to onset of patient reported functional decline, an important milestone in clinical progression.^7^

We aimed to describe clinical and biological characteristics of phenoconverters and to assess biological congruency among individuals with iRBD, hyposmia, NMCs, and healthy controls (HC) enrolled in PPMI. We tested two hypotheses: (1) that clinical phenoconversion does not necessarily align with biomarkers of synucleinopathy or dopaminergic deficit, and (2) that phenoconversion might often occur later than onset of meaningful functional decline.

## Methods

### Study population

This is a case series of individuals enrolled in PPMI, a prospective, international, multi-center cohort study (ClinicalTrials.gov number NCT01141023).^12^ Detailed information about inclusion criteria, informed consent, demographic data, and the study design can be found on the PPMI web site (http://www.ppmi-info.org/). We performed a descriptive analysis of participants enrolled across prodromal and HC cohorts. Prodromal groups included individuals with iRBD (confirmed either by overnight video-polysomnography or clinical diagnosis (some of the latter also requiring hyposmia and DAT abnormalities for enrollment)), hyposmia (based on University of Pennsylvania Smell Identification Test (UPSIT) ≤ 15th percentile applying age– and sex-adjusted norms; having an abnormal DAT was also required for the majority (75-80%) of the hyposmia group), and NMC of genetic variants associated with PD, including *GBA1, LRRK2*, and *SNCA*. Functional impairment was assessed by the anchors as proposed in NSD-ISS.^7^

### Inclusion and exclusion criteria

Participants were included if they: 1) belonged to a prodromal enrollment groups (iRBD, hyposmia), carried genetic variants associated with PD risk (*LRRK2, GBA1, SNCA*), or were HCs, 2) developed a new diagnosis (PD, DLB, MSA, PSP, and Alzheimer’s disease (AD) or other dementias) assigned by the clinical investigator after baseline, 3) had CSFaSynSAA and DAT imaging within ± two years of phenoconversion, and 4) had ≥1 follow-up visit. We excluded phenoconverters whose final diagnosis reverted to a prodromal stage, as well as those with absence of follow-up after baseline. Participants with missing biological data were excluded. with predefined exception as follows. First, participants with positive CSFaSynSAA or DAT results obtained before phenoconversion were included irrespective of timing. Positive results obtained after phenoconversion were included only if performed within two years. Negative CSFaSynSAA or DAT results were included only when obtained within two years before or after phenoconversion (Figure 1).

**Figure 1.**
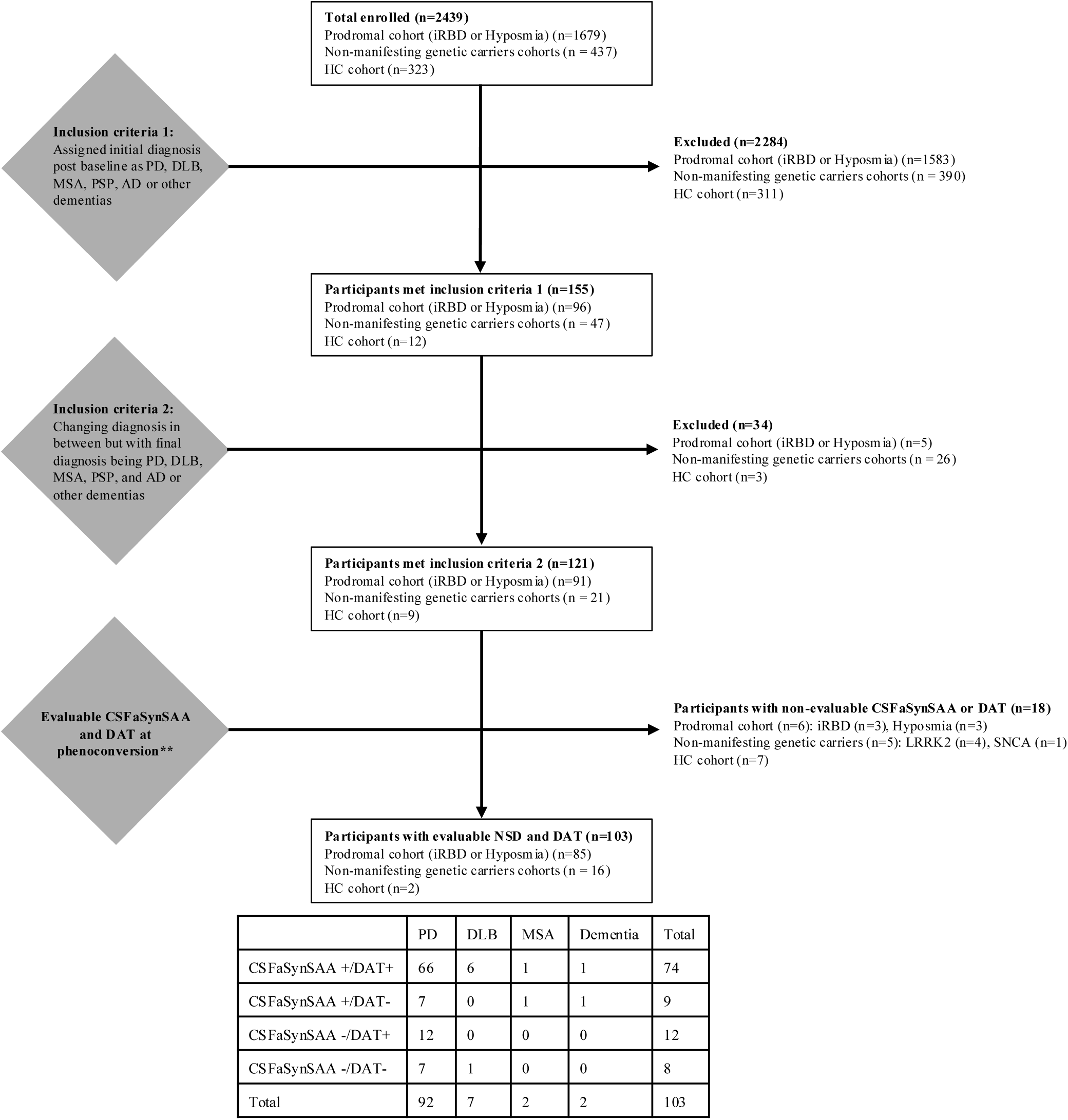
PPMI phenoconversion analysis data flowchart. Parkinson’s disease (PD), dementia with Lewy bodies (DLB), multiple system atrophy (MSA), Alzheimer’s disease (AD), α-synuclein pathology in CSF (CSFaSynSAA+), dopaminergic dysfunction imaging (dopamine transporter (DAT+). **If SAA or DAT was missing, positive results before PC were carried forward without time limit, positive results after PC were carried backward if within two years; negative results were carried forward or backward if within 2 years.

### Variables

Once enrolled at a site, PPMI participants underwent inDperson visits. Evaluations involved standardized questionnaires and clinician assessments covering both motor and nonDmotor symptoms, as well as neuropsychological testing as described in the supplement.^12^ Demographic variables included biological age, sex, age at phenoconversion and number of years of formal education. Clinical variables were based on Movement Disorder Society-Unified Parkinson’s Disease Rating Scale (MDS-UPDRS) Part I (non-motor experiences of daily living), Part II (motor aspects of experiences of daily living) and Part III (motor examination) total scores. Cognition was assessed with an objective cognitive test (Montreal Cognitive Assessment (MoCA)) and item 1 of the MDS-UPDRS Part I which asks participants for alterations in cognitive function, such as slow thinking, impaired reasoning, memory loss, and attention deficits. Smell test was another non-motor clinical variable, which was tested using the UPSIT. Diagnosis of dementia was based on investigator diagnosis.

### NSD-ISS anchors

NSD–ISS proposed a biologically anchored staging framework integrating biologically defined diagnosis and staging with increasing degree of functional impairment. It comprises six stages: stage 0 (NMC of fully penetrant pathogenic *SNCA* variant), stage 1A/B (CSFaSynSAA+ without (1A) or with dopaminergic dysfunction (D+) (1B), without clinical manifestations), stage 2A/B (CSFaSynSAA+ with clinical manifestations but no functional impairment), stage 3-6 (biomarker abnormalities and progressive functional impairment).^13^ The motor domain is defined by having subthreshold parkinsonism (MDS-UPDRS Part III score ≥ 5, excluding the postural and action tremor items). Non-motor symptoms are based on hyposmia, iRBD and cognitive impairment. Cognitive impairment is defined by MoCA total score ≤ 24 and MDS-UPDRS 1.1 ≥ 1. The clinical anchors are based on progressive functional impairment driven by motor, cognitive, and other non-motor domains as described.^14,15^

Biomarkers were CSFaSynSAA and DAT. CSFaSynSAA testing was conducted at Amprion.^6^ Briefly, the CSFaSynSAA had a dual output: 1 for the detection or notDdetection of α-synuclein SAA seeds (positive, negative, and inconclusive) and another for the type of seeds detected (Type I, Type II, and undetermined). Positive samples with Type I seeds presented high fluorescence values (≥45,000RFU) and these seeds are predominantly found in participants with NSD, whereas Type II seeds presented intermediate fluorescence values (≥3,000RFU and <45,000RFU) and these seeds are predominantly found in patients with underlying glial cytoplasmic inclusions or clinical presentation of MSA. Two assays were used to generate CSFaSynSAA results at the time of phenoconversion: the 150-hour assay and the 24-hour assay. The former does not distinguish between Type I and Type II, whereas the latter does.

DAT results are reported as the specific binding ratio (SBR) and as reflecting dopaminergic dysfunction (yes/no) based on an SBR value < 75% of the age– and sex-expected lowest putamen.

### Biological congruency definition close to phenoconversion

We defined biological congruency and incongruency based on CSFaSynSAA and DAT imaging profiles in relation to clinical diagnosis and genotype close to phenoconversion. We considered participants to be biologically “congruent” if they had biomarkers signature of CSFaSynSAA+/DAT+ specifically if they phenoconverted to PD. Participants were classified as biologically “incongruent” when their biological profile did not match the expected pattern for their clinical trajectory. This included individuals who converted to PD but lacked abnormal CSFaSynSAA or DAT findings, with two exceptions. For *LRRK2* NMC, biological congruency was driven solely by the presence of a DAT+ as *LRRK2*-related PD is known to potentially be associated with lack of CSFaSynSAA pathology in about 30% of cases.^9^ For people who converted to DLB, biological congruency was based solely on CSFaSynSAA+ regardless of DAT, as 3% to 9% of early DLB cases have normal DAT.^16,17^

### Statistical analysis

We performed descriptive analyses to summarize demographic, clinical, and biological characteristics close to phenoconversion, as well as preceding phenoconversion, stratified by enrolment cohort and subgroups. Categorical variables are reported as frequencies (percentages), and continuous variables are summarized using the mean and standard deviation (SD) or median and interquartile range (IQR), as appropriate.

Statistical analyses were conducted using SAS version 9.4 (SAS Institute Inc., Cary, NC). Data visualizations were performed using RStudio (Boston, MA;) with R Vienna, Austria) and the ggplot2 package (https://ggplot2.tidyverse.org).

Data used were obtained on 21-January-2025 from www.ppmi-info.org/access-data-specimens/download-data (RRID:SCR 006431), with exception of CSFaSynSAA data (obtained on 9-June-2025). Statistical analysis codes used to perform the analyses in this article are shared on Zenodo (10.5281/zenodo.17992083).

## Results

There were 121 phenoconverters across all groups. The number of phenoconversion events differed substantially across groups. The iRBD group had the highest annualized phenoconversion rate amongst all groups (7.9%), followed by hyposmia (4.2%). The NMCs had annual rate of phenoconversion below 2% similar to our HC group (*LRRK2*: 1.3%, *GBA1*: 0.3%, *LRRK2*+*GBA1*: 0.9%, HC: 0.5%) (Table S1). All *GBA1* NMCS had N409S variant, and *LRRK2* NMCs had G2019S.

Of the 121 phenoconverters, 103 had available biomarker data within ± two years from phenoconversion and were included in further analysis. None of the *SNCA* NMCs had available biomarkers within ± 2 years (Figure 1). Clinical characteristics close to phenoconversion are presented in Table 1. PD was the most common diagnosis in all prodromal groups (78.7% iRBD, 100% hyposmia, 100% *GBA1* and *LRRK2* NMC). The dual *LRRK2/GBA1* NMCs also converted to PD. DLB was observed exclusively in iRBD (14.9%), while a small proportion of iRBD (4.3%) were diagnosed as MSA. Two HC participants also phenoconverted, one to PD and one to AD.

**Table 1.** Characteristics at the time of phenoconversion of participants with evaluable CSFaSynSAA and DAT (N=103)

| Variables <sup>a</sup> | HC<br>(N = 2) | iRBD <sup>d</sup><br>(N = 47) | Hyposmia<br>(N = 38) | *GBA1*<br>(N = 3) | *LRRK2*<br>(N = 12) | *LRRK2+GBA1*<br>(N = 1) |
| --- | --- | --- | --- | --- | --- | --- |
| Age, median<br>(IQR) | 80.2<br>(79, 81) | 70.0<br>(67, 76) | 72.6<br>(68, 78) | 71.2<br>(69, 73) | 67.3<br>(59, 78) | 67.6<br>(68, 68) |
| Male, n (%) | 1 | 34 (72.3%) | 28 (73.7%) | 2 | 7 (58.3%) | 0 |
| Primary research diagnosis <sup>b</sup>, n (%) |  |  |  |  |  |  |
| Idiopathic PD | 1 | 37 (78.7%) | 38 (100.0%) | 3 | 12 (100.0%) | 1 |
| Dementia with Lewy bodies | 0 | 7 (14.9%) | 0 | 0 | 0 | 0 |
| Multiple system atrophy | 0 | 2 (4.3%) | 0 | 0 | 0 | 0 |
| Other Dementia | 1 | 1 (2.1%) | 0 | 0 | 0 | 0 |
| Months from baseline to PC, Median<br>(IQR) | 31.2<br>(14, 49) | 13.5<br>(11, 28) | 13.0<br>(7, 28) | 35.9<br>(29, 66) | 46.6<br>(12, 61) | 84.9<br>(85, 85) |
| CSFaSynSAA status, n (%) |  |  |  |  |  |  |
| Negative | 0 | 9 (19.1%) | 1 (2.6%) | 2 | 8 (66.7%) | 0 |
| Positive Type 1 | 2 | 38 (80.9%) | 37 (97.4%) | 1 | 4 (33.3%) | 1 |
| Positive Type 2 | 0 | 0 | 0 | 0 | 0 | 0 |
| NSD-ISS stage (NSD subset only), n (%) |  |  |  |  |  |  |
| 2A | 1 | 4 (10.5%) | 4 (10.8%) | 0 | 0 | 0 |
| 2B | 1 | 3 (7.9%) | 9 (24.3%) | 1 | 2 (50.0%) | 1 |
| 3 | 0 | 16 (42.1%) | 17 (45.9%) | 0 | 2 (50.0%) | 0 |
| 4 | 0 | 13 (34.2%) | 7 (18.9%) | 0 | 0 | 0 |
| 5 | 0 | 1 (2.6%) | 0 | 0 | 0 | 0 |
| 6 | 0 | 1 (2.6%) | 0 | 0 | 0 | 0 |
| MDS-UPDRS-III, median<br>(IQR) | 19.0<br>(4, 34) | 13.0<br>(6, 22) | 12.0<br>(8, 18) | 13.0<br>(7, 22) | 10.0<br>(6, 12) | - |
| MDS-UPDRS-III change, median<br>(IQR) | 6.0<br>(4, 8) | 6.0<br>(1, 12) | 5.5<br>(2, 12) | 5.5<br>(3, 8) | 3.5<br>(2, 7) | - |
| Subthreshold parkinsonism <sup>c</sup> | 1 | 37 (78.7%) | 30 (85.7%) | 3 | 8 (72.7%) | 1 |
| MDS-UPDRS-II, median (IQR) | 1.5 (0, 3) | 5.0 (3, 10) | 6.0 (3, 9) | 1.0 (0, 5) | 3.0 (0, 9) | 1.0 (1, 1) |
| MoCA, n (%) |  |  |  |  |  |  |
| • ≥ 26 | 1 | 26 (55.3%) | 22 (59.5%) | 2 | 8 (72.7%) | 1 |
| • 21 - 25 | 1 | 14 (29.8%) | 14 (37.8%) | 1 | 3 (27.3%) | 0 |
| • < 21 | 0 | 7 (14.9%) | 1 (2.7%) | 0 | 0 | 0 |
| MoCA change, median (IQR) | 0.5 (-2, 3) | 0.0 (-2, 2) | -0.5 (-1, 1) | -0.5 (-1, 0) | 1.0 (0, 2) | 1.0 (1, 1) |

(a) Missing data: MoCA:1 Hyposmia, 1 *LRRK2*; MoCA change: 12 iRBD, 16 hyposmia, 1 *GBA1*, 3 *LRRK2*; MDS-UPDRS-III: 3 hyposmia, *LRRK2*: 1; MDS-UPDRS-III change: 12 iRBD, 18 hyposmics, 1 *GBA1*, 4 *LRRK2*. (b) One of the RBD participants was diagnosed with both PD and Dementia; (c) Subthreshold parkinsonism is defined as MDS-UPDRS-III $\geq 5$ , excluding postural and action tremor. (d) 36/47 (76.6%) were PSG-confirmed RBD and 1 with missing PSG data. PC: phenoconversion. Note: Percentages may not total 100% due to rounding.

Median (IQR) time to phenoconversion differed by group, with 13.5 months (11–28) in the iRBD group, 13.0 months (7–28) in the hyposmia group, and the longest median interval observed in the *LRRK2* group at 46.6 months (12–61) (Table 1).

### Clinical characteristics close to phenoconversion

Demographic and clinical characteristics of participants close to phenoconversion are presented in Table 1. iRBD and hyposmic phenoconverters showed similar age distributions (median (IQR) 70.0 years (67–76) and 72.6 years (68–78), respectively), and both groups were predominantly male (iRBD males: 72.3% and hyposmic males: 73.7%). *LRRK2*-NMCs phenoconverters were slightly younger (median (IQR) 67.3 years (59–78)), while HC phenoconverters were considerably older (median (IQR) 80.2 years (79–81)) with similar sex distribution (*LRRK2*: 58.3% males vs HC: 50% males).

Motor impairment at phenoconversion was common across all groups. Most phenoconverters met criteria for subthreshold parkinsonism regardless of final diagnosis (Table 1). Cognitive status showed greater heterogeneity amongst groups. While phenoconverters commonly had normal cognitive scores, over 40% of iRBD phenoconverters had investigator determined either mild cognitive impairment or dementia at time of phenoconversion. Mild cognitive impairment was also noted in nearly 38% hyposmics, and one had dementia. The majority of NMCs had a normal cognitive assessment and none had dementia (Table 1).

MDS-UPDRS-III scores worsened across all groups in the year prior to phenoconversion, with median annual increases ranging between 4 and 6 points, indicating progressive motor deterioration shortly prior to clinical diagnosis. In contrast, MoCA scores showed minimal change over the same interval (Table 1).

### Biomarkers at phenoconversion

We evaluated CSFaSynSAA and DAT imaging biomarkers within ± two years from phenoconversion (Table 2). Most prodromal phenoconverters were CSFaSynSAA+: 38/47 (80.8%) iRBD and even higher, 38 (97.4%), among hyposmics. In contrast, a higher proportion than expected (66.7% (8/12)) *LRRK2*-NMCs phenoconverters were CSFaSynSAA–. Of note, 83% of *LRKK2*-NMCs were normosmics (Table S2). Among *GBA1*-NMCs, only 1/3 was CSFaSynSAA+. Both HC, one who phenoconverted to AD and one to PD, were CSFaSynSAA+. None of the phenoconverters showed type II CSFaSynSAA characteristics.

**Table 2.** Biological characterization of all phenoconverters close to phenoconversion (within two years) (N=103)

| **PD/DLB/MSA**<br>**/Other Dementia** | **HC**<br>**(N = 2)** | **iRBD**<br>**(N = 47)** | **Hyposmia**<br>**(N = 38)** | ***GBA1***<br>**(N = 3)** | ***LRRK2***<br>**(N = 12)** | ***LRRK2+GBA1***<br>**(N = 1)** |
| --- | --- | --- | --- | --- | --- | --- |
| CSFaSynSAA+ <sup>1</sup>/DAT+<br>(N = 74) | 1 | 34 (72.3%) | 33 (86.8%) | 1 | 4 (33.3%) | 1 |
| CSFaSynSAA+ <sup>1</sup>/DAT-<br>(N = 9) | 1 | 4 (8.5%) | 4 (10.5%) | 0 | 0 | 0 |
| CSFaSynSAA- <sup>2</sup>/DAT+<br>(N = 12) | 0 | 6 (12.8%) | 1 (2.6%) | 0 | 5 (41.7%) | 0 |
| CSFaSynSAA- <sup>2</sup>/DAT-<br>(N = 8) | 0 | 3 (6.4%) | 0 | 2 | 3 (25.0%) | 0 |

(1) Amongst 83 CSFaSynSAA+: 21 had a 150-hour assay and 62 had 24-hour assay. (2) Amongst 20 CSFaSynSAA-, 4 had 150-hour assay and 16 had 24-hour assay. All of these 24-hour results were concordant with the 150-hour assay results. Note: Percentages may not total 100% due to rounding.

DAT abnormalities were more frequently observed in the hyposmia group, while more heterogeneous in iRBD. Overall, amongst the 92 participants who phenoconverted to PD, 14 (15.2%) could be considered subjects without dopamine deficit (SWEDDs) as they had a normal DAT imaging close to phenoconversion (Table S3). The proportion of DAT+ varied by group: 86.5% (32/37) in iRBD, 89.5% (34/38) in hyposmia, 33.3% (1/3) in *GBA1* NMCs, and 75% (9/12) in *LRRK2* NMCs (Table S3). It is worth noting that 75-80% of hyposmia participants were required to have abnormal DAT binding at enrollment while individuals with PSG confirmed iRBD were allowed to be recruited independent of DAT deficit.

When combining CSFaSynSAA and DAT results, a high proportion of phenoconverters in the iRBD and hyposmia groups demonstrated biologically congruent profiles (Figure 2). Using CSFaSynSAA+/DAT+ as the canonical biomarker profile seen in clinically manifest synucleinopathies, 72.3% of iRBD (34/47) and 86.8% of hyposmia phenoconverters (33/38) fell within this biologically aligned category (Table 2). In contrast, biological heterogeneity was more frequent in genetic groups. Among *LRRK2* phenoconverters, 41.7% (5/12) showed an CSFaSynSAA−/DAT+ pattern, consistent with the known heterogeneity of *LRRK2*-associated proteinopathies. Notably, 25% (3/12) were CSFaSynSAA−/DAT− despite a clinical diagnosis of PD. Unexpected biological profiles (“biological incongruency”) were also observed in *GBA1* NMC: 66.7% (2/3) of *GBA1* phenoconverters to PD demonstrated CSFaSynSAA–/DAT– patterns, which is not expected in *GBA1*-PD synucleinopathy.

**Figure 2.**
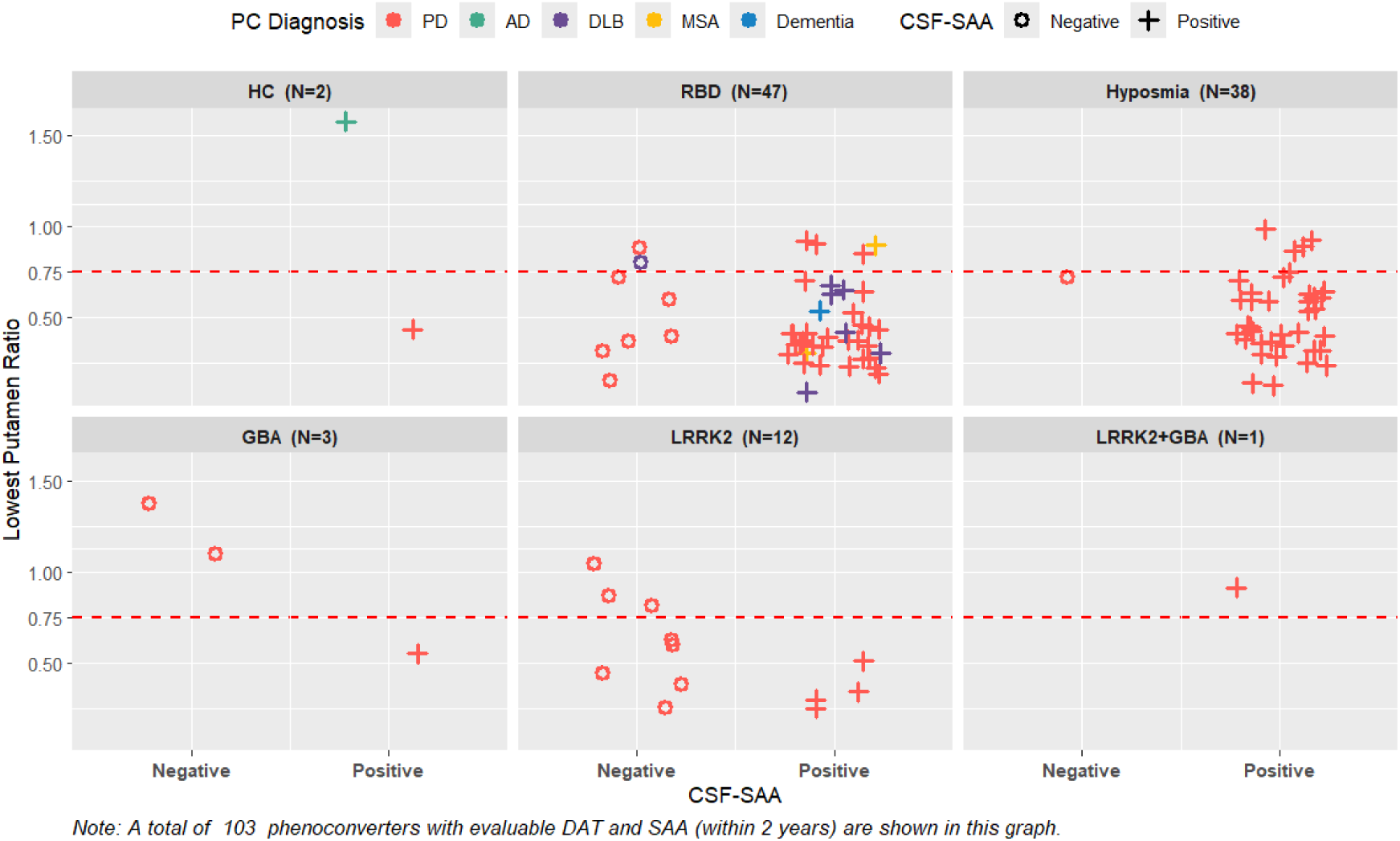
DAT vs CSFaSynSAA close to phenoconversion (N=103)

All phenoconverters to DLB were in the iRBD group. Six of seven DLB phenoconverters showed biologically congruent profiles (CSFaSynSAA+/DAT+), with one case having both negative biomarkers. Phenoconversion to MSA was observed in two iRBD cases, both of whom tested positive for CSFaSynSAA type I, with no cases having type II CSFaSynSAA. DAT results were variable, with one case testing positive and the other remaining negative.

### Functional NSD staging at phenoconversion

NSD-ISS staging close to phenoconversion is presented in Figure 3. The majority were NSD stage 3 (CSFaSynSAA+/DAT+ with slight functional impairment), and consistent with a new clinical diagnosis; however, a wide range of stages was observed. In iRBD, 42.1% were stage 3, 34.2% stage 4, 2.6% stage 5 and 2.6% stage 6 at phenoconversion. Hyposmic phenoconverters similarly clustered in stages 2B–3–4 (2B: 24.3%, 3: 45.9%, 4: 18.9%). In contrast, the four *LRRK2* cases were evenly split across stages 2B and 3 (Figure 3).

**Figure 3.**
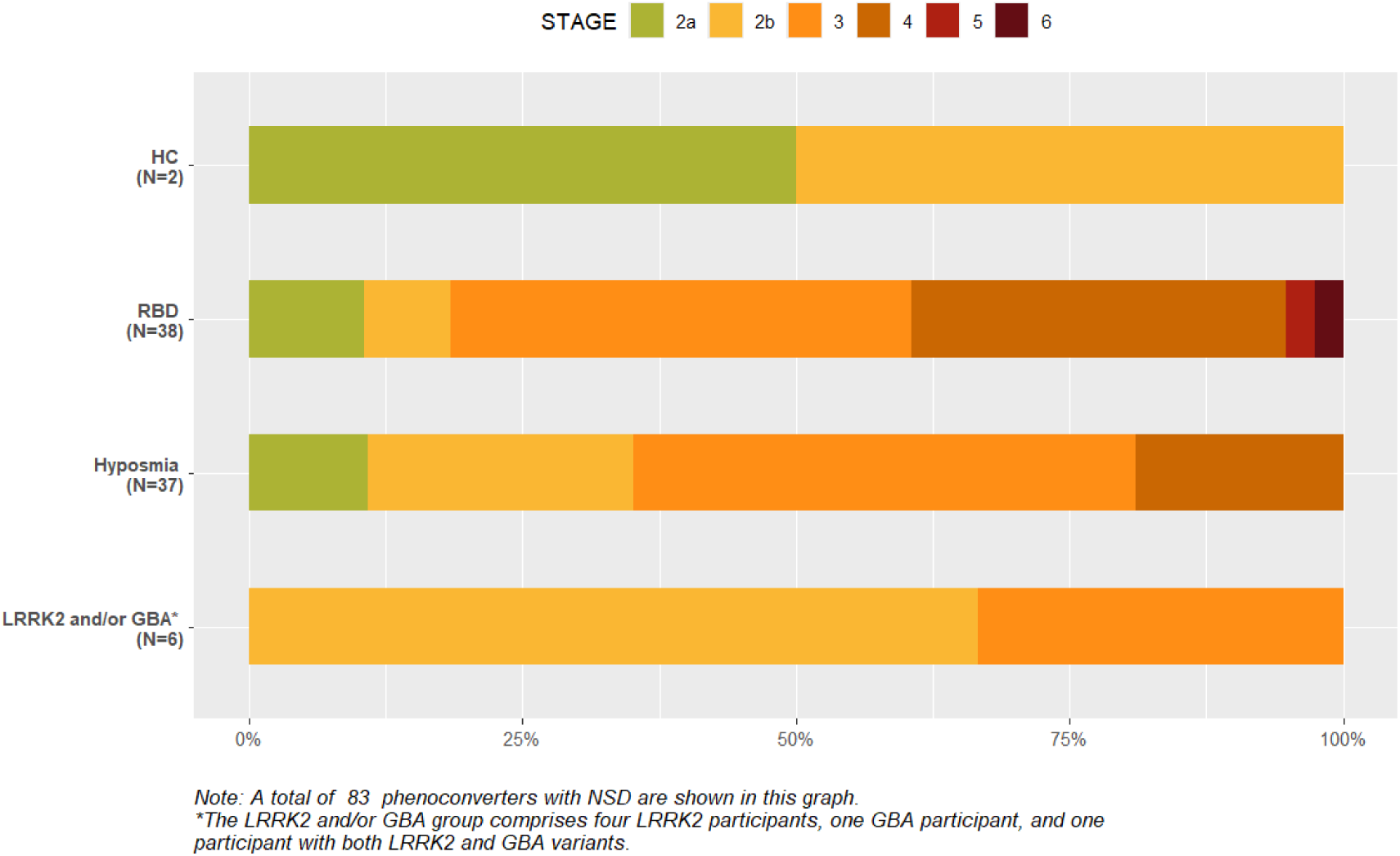
Stage by enrolment group close to initial phenoconversion (N=83)

We further examined domains of functional impairment in individuals with NSD-ISS ≥4. At phenoconversion, 15 individuals with iRBD and 7 with hyposmia met NSD-ISS ≥4 criteria. While in hyposmic participants, functional impairment appeared to be primarily driven by other non-motor symptoms, in the iRBD group there were higher rates of both cognitive and other non-motor symptoms, suggesting a more extensive and multisystem disease burden at the time of phenoconversion (Table 3).

**Table 3.**
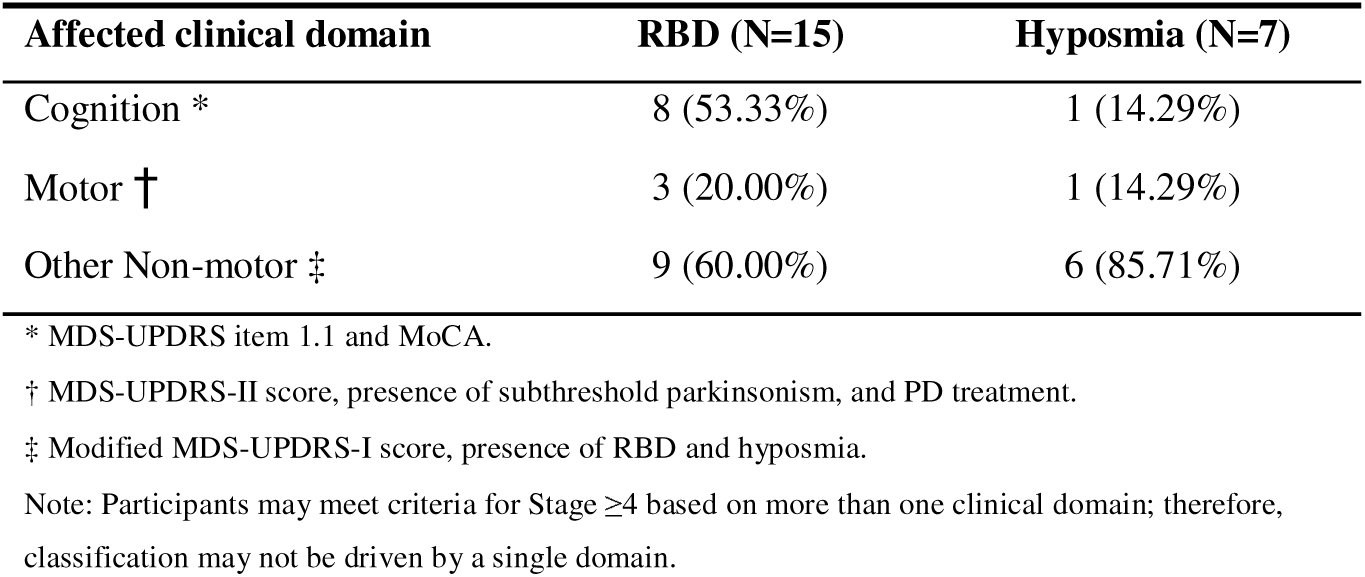
NSD-ISS Clinical domain status for phenoconverters at NSD-ISS ≥4.

| **Affected clinical domain** | **RBD (N=15)** | **Hyposmia (N=7)** |
| --- | --- | --- |
| Cognition * | 8 (53.33%) | 1 (14.29%) |
| Motor † | 3 (20.00%) | 1 (14.29%) |
| Other Non-motor ‡ | 9 (60.00%) | 6 (85.71%) |

\* MDS-UPDRS item 1.1 and MoCA.
† MDS-UPDRS-II score, presence of subthreshold parkinsonism, and PD treatment.
‡ Modified MDS-UPDRS-I score, presence of RBD and hyposmia.
Note: Participants may meet criteria for Stage $\geq 4$ based on more than one clinical domain; therefore, classification may not be driven by a single domain.

## Discussion

In this study we examined clinical and biomarker characteristics close to phenoconversion across four at-risk groups (i.e., iRBD, hyposmia, and *LRRK2* and *GBA1*-NMCs) from study enrollment until phenoconversion to a clinically-defined neurodegenerative disorder. The novelty of our data is availability of biomarker characterization of individuals close to phenoconversion which has not been explored systematically before across these distinct prodromal groups. This study provides several novel insights. First, the explicit categorization of “biologically congruent” versus “biologically incongruent” phenoconverters is a new conceptual framework that directly tests alignment between clinical diagnosis and biology. The identification of a sizeable subgroup of clinically diagnosed individuals who lacked biological evidence of synucleinopathy or dopaminergic deficit, specifically with the diagnosis of PD, challenges the assumption that clinically-defined phenoconversion without biological characterization is a reliable milestone in disease continuum. Second, the use of functional staging within the NSD−ISS to contextualize phenoconversion revealed a systematic delay between biological/functional change and clinical diagnostic assignment, a phenomenon not previously quantified. While prior studies reported that motor and cognitive decline begins before PD diagnosis, our work integrates these trajectories with biomarker-defined biological staging.^18^ Third, our analysis suggests potential under-recognition of prodromal DLB within iRBD groups. Although this has been speculated, our data show that cognitive markers declined prior to diagnosis in cases ultimately classified as PD, indicating possible misdiagnosis or insufficient use of supportive cognitive tools. The latter finding might be different if the focus of the cohort was on prodromal DLB features.

The hyposmia group displayed the most homogeneous profile, with the majority of individuals (87%) being CSFaSynSAA+/DAT+. This is consistent with prior PPMI analyses showing that olfactory impairment strongly correlates with CSFaSynSAA+ and DAT deficit predicting future risk of PD.^9,19,20^ However, more data, and ideally quantitative measures of CSFaSynSAA and DAT (beyond single threshold), are required to reliably estimate the expected timeline to development of clinical phenotype. Similarly, a substantial proportion (72%) of iRBD individuals were CSFaSynSAA+/DAT+. In contrast, one quarter of iRBD phenoconverters displayed a biologically unexpected profile (four DAT−, six CSFaSynSAA−, and three with both negative) (Table 2). While an argument can be made that these participants are close to DAT deficit threshold used in PPMI, they meet criteria for SWEDD, and prior longitudinal data demonstrated lack of longitudinal progression in SWEDDs.^21^ Both concepts will need to be tested with a longer follow up.

In *LRRK2* NMC, CSFaSynSAA heterogeneity was expected.^22,23^ Consistent with prior literature nearly 30-35% of *LRRK2* phenoconverters were CSFaSynSAA–, though in our cohort that was even higher.^9^ Accounting for expected proteinopathy heterogeneity, three quarters of *LRRK2* (4/12 CSFaSynSAA+/DAT+ and 5/12 CSFaSynSAA–/DAT+) were biologically “congruent”. CSFaSynSAA negativity was higher than expected among *LRRK2* phenoconverters (67%), which could be explained by mixed underlying pathology. This interpretation is supported by a high proportion of normosmics (83%) amongst *LRRK2* NMCs, along with a notable proportion of females (42%), both factors previously associated with lower CSFaSynSAA positivity.^24^ Three out of 12 were CSFaSynSAA–/DAT– despite phenoconversion diagnosis of PD. The number of *GBA1* phenoconverters was exceedingly low (N=3), consistent with the reduced penetrance of *GBA1* variants (i.e., low probability that NMC will develop PD during their lifetime). This is particularly relevant for the N409S variant, which was predominant in our cohort, though biological characteristics were unexpected. Two out of three *GBA1* NMCs were CSFaSynSAA– which contradicts established data that *GBA1* is a pure synucleinopathy. In any case, very low conversion rates and extended time to phenoconversion in *GBA1* and *LRRK2* NMC indicate that targeting these groups for therapeutic trials is not yet feasible unless biological profiles predictive of progression are used for enrichment. Without such enrichment strategies, unselected cohorts would require impractically large sample sizes and excessively long follow-up periods.

Beyond biological characterization, our analysis of functional staging showed that many individuals had already reached NSD-ISS stage ≥4, indicating mild but meaningful functional impairment, by the time phenoconversion was assigned. Motor trajectories similarly showed clear deterioration during the year preceding diagnosis. Cumulatively, these findings suggest that functional impairment NSD-ISS definitions may be more sensitive markers of early disease progression than canonical phenoconversion, which appears to occur later in the biological and clinical continuum.

It is important to note that phenoconversion was defined within a research context and based on investigator-assigned clinical diagnosis. As such, it is likely that it captures the onset of diagnostic features earlier than would occur in routine clinical practice. In real-world scenarios, symptom onset is usually first recognized by the patient, followed by primary care doctor who refers them to a neurologist to make the final diagnosis. This multistep pathway inevitably introduces diagnostic delay. Consequently, the interval observed here between phenoconversion and meaningful functional decline is likely an underestimate of the delay that occurs in routine care, where phenoconversion would typically be identified at a later stage of disease progression.

The rates of phenoconversion varied widely across the PPMI prodromal cohorts, with iRBD showing the highest conversion frequencies, followed by hyposmia, while *GBA1* and *LRRK2* groups exhibited very low rates and long latencies to phenoconversion. Our findings reinforce observations previously reported in prodromal PD research.^25–30^ iRBD is recognized as the strongest prodromal marker of synucleinopathy, with conversion rates of 30–90% over 10–15 years.^15,31^ However, these cohorts were not biomarker characterized. In our iRBD group, we found a relatively low proportion of DLB phenoconverters DLB (7/47). This contrasts with other studies.^32^ This difference may be explained by 30% of them being CSFaSynSAA–, recruitment source (movement disorders vs cognitive clinics) and/or under-recognition of early cognitive impairment, early visuospatial impairment or subtle cognitive fluctuations by site investigators. However, previously reported cohorts were not systematically biomarkers characterized so more longitudinal data in biomarkers defined cohorts are necessary. Two participants with iRBD phenoconverted to MSA, neither of whom had MSA-type CSFaSynSAA (type II). One case with MSA was CSFaSynSAA Type I+/DAT−, raising uncertainty regarding DAT thresholds in MSA phenotypes. This finding also suggests the possibility of a cerebellar type of MSA as it has been described as having relatively preserved nigrostriatal dopaminergic function, with normal or near-normal DAT imaging, in keeping with current MDS diagnostic criteria.^33^

Several limitations must be acknowledged. This was an observational and descriptive study and does not establish causality. The timing of biomarker acquisition varied across individuals. Although we restricted analyses to biomarkers assessed within ± two years of phenoconversion, biological change may have occurred earlier or later. The reliance on investigator-assigned diagnoses may introduce site-level variability and be subject to misclassification. This is important in biologically congruent individuals given the short follow-up which could be explained by incorrect diagnoses or distinct underlying biology. Additionally, heterogeneity across groups limits direct comparability: genetic cohorts were small, enrolled earlier, at younger age, and largely without biomarker enrichment, representing a fundamentally different population from iRBD and hyposmic participants. The majority of hyposmia group was enriched by having an abnormal DAT by the time of inclusion in the study which might have caused a selection bias. CSFaSynSAA and DAT measures were not always contemporaneous. The NSD-ISS staging framework has not yet been fully validated across diverse prodromal populations, which limits its generalizability at very early disease stages. In this context, commonly used cognitive screening tools such as the MoCA may lack sensitivity for detecting early cognitive changes in prodromal DLB. Finally, the low conversion rates in genetic groups (especially *GBA1*) limit statistical power for subgroup analyses.

To conclude, phenoconversion may be non-congruent and temporally delayed relative to both biological changes and functional impairment, raising concerns about its reliability as an outcome measure in clinical trials aiming to intervene prior to traditional clinical diagnosis. Cumulatively, these findings highlight the need for refined, biologically anchored definitions of disease progression that incorporate at minimum biomarkers of synuclein pathology and dopaminergic dysfunction, and functional staging. Integrating these domains into future frameworks could improve the sensitivity and specificity of early diagnostic criteria and enhance trial design for disease-modifying therapies improving participant selection. Armamentarium of biomarkers will expand to allow more comprehensive characterization of underlying disease pathobiology.

## Supporting information

Supplementary Table S1. Annualized phenoconversion rate by cohort

Supplementary Table S2. University of Pennsylvania Smell Identification Test (UPSIT) results at the time of phenoconversion

Supplementary Table S3. Biological characterization of PD phenoconverters at the time of phenoconversion

## Data Availability

All data produced are available online at https://www.ppmi-info.org/access-data-specimens/download-data

## Acknowledgment

We are grateful to all PPMI participants and their families for their invaluable contribution to the PPMI. We also thank the investigators, coordinators, and staff across all participating PPMI sites for their dedication to data collection and study management.

PPMI – a public-private partnership – is funded by the Michael J. Fox Foundation for Parkinson’s Research and funding partners, including Abbvie, Alamar Biosciences, Aligning Science Across Parkinson’s, Arrowhead Pharma, AskBio, BIAL, BioArctic, Biohaven, BlueRock Therapeutics, Bristol-MyersSquibb, Calico Labs, Capsida Biotherapeutics, Critical Path Institute, DaCapo Brainscience, Denali, Edmond J. Safra Foundation, Eli Lilly, Gain Therapeutics, GE HealthCare, Genentech, GSK, Insitro, Johnson & Johnson Innovative Medicine, Lundbeck, Merck, Neumora, Neuron23, Novartis, Regeneron, Roche, Sanofi, Tenvie, UCB, VanquaBio, Voyager Therapeutics, the Weston Family Foundation.

Protocol information for The Parkinson’s Progression Markers Initiative (PPMI) Clinical – Establishing a Deeply Phenotyped PD Cohort AM 3.2. can be found on protocols.io or by following this link: https://dx.doi.org/10.17504/protocols.io.n92ldmw6ol5b/v2.

## Authors’ Roles

Clarification of role(s): design (D), execution (Ex), analysis (A), writing (W), editing (Ed) of final version of the manuscript.

CS: D, Ex, A, W, Ed

JY: A, W, Ed

LC: D, A, Ed

DW: D, Ed

KC: A, Ed

CCG: A, Ed

DEL: A, Ed

AN: D, Ed

AS: D, Ed

CT: D, Ed

EB: D, Ed

TFT: D, Ed

BM: D, Ed

RNA: D, Ed

KP: D, Ed

KM: D, Ed

TS: D, A, Ed

## Financial Disclosure/Conflict of Interest concerning the research related to the manuscript

All information on support and financial issues from all authors relevant to the research covered in the submitted manuscript must be disclosed, regardless of date. Additional financial information unrelated to the current research covering the past year will also be disclosed as part of a completed ICMJE form (see below).

**Funding Sources for study.**

## Tables and figures

- Table 1. Characteristics at the time of phenoconversion of participants with evaluable CSFaSynSAA and DAT (N=103)
- Table 2. Biological characterization of all phenoconverters close to phenoconversion (within two years) (N=103)
- Table 3. Clinical domain status for phenoconverters at NSD-ISS ≥4
- Figure 1. Flow chart of selection criteria (N = 103)
- Figure 2. DAT vs CSFaSynSAA close to phenoconversion (N=103)
- Figure 3. Stage by enrolment group close to initial phenoconversion (N=83)

