## Supplementary Table S1. Annualized phenoconversion rate by cohort for "Synuclein and dopamine transporter biomarkers among phenoconverters to parkinsonian disorders"

| **Table S1. Annualized phenoconversion rate by cohort (N = 121)** | | | | | |
| --- | --- | --- | --- | --- | --- |
| **Cohort Subgroup** | **N** | **Events (Phenoconverted)** | **Total Person-years** | **Events per year** | **Annualized PC rate (% year)** |
| HC | 290 | 9 | 1862.2904 | 0.005 | 0.5% |
| RBD | 378 | 50 | 633.24384 | 0.079 | 7.9% |
| Hyposmia | 809 | 41 | 985.63014 | 0.042 | 4.2% |
| *GBA1* | 167 | 3 | 1033.9918 | 0.003 | 0.3% |
| *LRRK2* | 191 | 16 | 1210.8411 | 0.013 | 1.3% |
| *LRRK2+GBA1* | 17 | 1 | 109.09315 | 0.009 | 0.9% |
| *SNCA* | 11 | 1 | 54.27123 | 0.018 | 1.8% |
