## Supplementary Table S2. University of Pennsylvania Smell Identification Test (UPSIT) results at the time of phenoconversion for "Synuclein and dopamine transporter biomarkers among phenoconverters to parkinsonian disorders"

| **Table S2. University of Pennsylvania Smell Identification Test (UPSIT) results at the time of phenoconversion** | | | | | | |
| --- | --- | --- | --- | --- | --- | --- |
|  | **HC (N = 2)** | **RBD (N = 47)** | **Hyposmia (N = 38)** | ***GBA1* (N = 3)** | ***LRRK2* (N = 12)** | ***LRRK2+GBA1* (N = 1)** |
| UPSIT Percentile ^a^ | | | | | | |
| Mean (SD) | 17.5 (16.3) | 13.9 (20.9) | 5.6 (3.9) | 47.2 (41.8) | 48.2 (28.5) | 4.0 (NA) |
| Median (IQR) | 17.5 (6, 29) | 5.0 (3, 12) | 5.0 (2, 8) | 37.5 (11, 93) | 55.5 (22, 70) | 4.0 (4, 4) |
| Range | (6, 29) | (1, 81) | (1, 17) | (11, 93) | (3, 84) | (4, 4) |
| Missing | 0 | 2 | 0 | 0 | 0 | 0 |
| UPSIT Percentile <= 15 ^a^ | | | | | | |
| No | 1 | 10 (22.2%) | 1^b^ (2.6%) | 2 | 10 (83.3%) | 0 |
| Yes | 1 | 35 (77.8%) | 37 (97.4%) | 1 | 2 (16.7%) | 1 |
| Missing | 0 | 2 | 0 | 0 | 0 | 0 |
| ^a^ Last observation carried forward.  ^b^ One participant met original inclusion criteria for the pre-2020 PPMI hyposmia cohort (enrolled 2013-2015) but exceeded the ≤15th-percentile cutoff (16.5) when recalculated using updated methodology (Brumm et al., 2023; <https://pubmed.ncbi.nlm.nih.gov/36849448/>). | | | | | | |
