## Supplementary Table S3. Biological characterization of PD phenoconverters at the time of phenoconversion for "Synuclein and dopamine transporter biomarkers among phenoconverters to parkinsonian disorders"

| **Table S3. Biological characterization of PD phenoconverters at the time of phenoconversion (N = 92)** | | | | | | |
| --- | --- | --- | --- | --- | --- | --- |
|  | **HC (N = 1)** | **RBD (N = 37)** | **Hyposmia (N = 38)** | ***GBA1* (N = 3)** | ***LRRK2* (N = 12)** | ***LRRK2+GBA1* (N = 1)** |
| CSFaSynSAA+/DAT+ ^1^ (N = 66) | 1 | 26 (70.3%) | 33 (86.8%) | 1 | 4 (33.3%) | 1 |
| CSFaSynSAA+/DAT- ^1^ (N = 7) | 0 | 3 (8.1%) | 4 (10.5%) | 0 | 0 | 0 |
| CSFaSynSAA-/DAT+ ^1^ (N = 12) | 0 | 6 (16.2%) | 1 (2.6%) | 0 | 5 (41.7%) | 0 |
| CSFaSynSAA-/DAT- ^1^ (N = 7) | 0 | 2 (5.4%) | 0 | 2 | 3 (25.0%) | 0 |

^1^ If SAA or DAT was not measured at the phenoconversion visit, positive results obtained before the visit were carried forward without time restriction, positive results obtained after the visit were carried backward if within two years, and negative results were carried forward or backward if obtained within two years.

Note: Percentages may not total 100% due to rounding.
